# Spatiotemporal dynamics, factors associated with neonatal mortality and health interventions in Mali: a modeling study

**DOI:** 10.64898/2026.09.17.26363290

**Authors:** Fatoumata Bintou Traoré, Cheick Sidya Sidibé, Youssouf Keita, Fatoumata Sidibé, Ibrahim Terera, Mariam Traoré, Mamadou Berthé, Haoua Dembelé, Boureyma Belem, Kassoum Koné, Aissata Touré, Abdoulaye S Dabo, Fatou Diawara, Ibrehima Guindo, Abdoulaye Maiga

**Affiliations:** National Public Health Institute, Bamako, Mali; National Training Institute in Health Sciences, Bamako, Mali; Global Financing Facility, the World Bank Group. Bamako Mali; University of Sciences, Techniques and Technologies of Bamako Mali; Johns Hopkins University Bloomberg School of Public Health, Baltimore, MD, USA

**Author notes:** Corresponding author: Fatoumata Bintou Traoré, MD, MPH, PhD.

## Abstract

**Background:** Neonatal mortality in Sub-Saharan Africa remains high, accounting for approximately 46% of all neonatal deaths worldwide. In Mali, neonatal mortality stands at 29 deaths per 1,000 live births, with marked subnational disparities. This study aimed to assess the spatiotemporal dynamics of neonatal mortality in Mali from 2012 to 2023 and to project potential reductions in neonatal mortality by 2035 under different scenarios of scaling up three high-impact interventions.

**Methods:** We analyzed data from the 2012-2013, 2018, and 2023-2024 Demographic and Health Surveys to assess spatiotemporal patterns in neonatal mortality across Mali. Multilevel mixed-effects logistic regression identified factors associated with neonatal mortality. Global Moran’s I and Local Indicators of Spatial Association assessed spatial dependence and identified high-high clusters. The Lives Saved Tool estimated potential reductions in neonatal deaths under alternative intervention scale-up scenarios through 2035.

**Results:** National neonatal mortality declined from 34 deaths per 1,000 live births (95% CI: 31-38) in 2012 to 29 (95% CI: 26-32) in 2023. Significant spatial clustering persisted across all survey rounds (Global Moran’s I, all p<0.001), with recurrent high-high clusters in Sikasso, Mopti, Segou and Tombouctou. Male neonates had higher odds of death (AOR, 1.66; 95% CI, 1.26-2.19), whereas birth intervals of at least 2 years (AOR range, 0.42-0.48), postnatal care (AOR, 0.42; 95% CI, 0.19-0.94), and improved sanitation (AOR, 0.42; 95% CI, 0.27-0.67) were associated with lower odds. LiST projections indicated that scaling up thermal care, neonatal resuscitation, and clean cord care could avert more than half of preventable neonatal deaths by 2035.

**Conclusions:** Persistent clusters of high neonatal mortality were identified in Sikasso, Segou, Mopti, and Tombouctou, highlighting the need for geographically targeted strategies. Scaling up high-impact neonatal interventions could substantially reduce preventable neonatal deaths in Mali.

## I. Introduction

Neonatal mortality is a significant concern in Mali as well as in sub-Saharan Africa, accounting for nearly 50% of under-5 child mortality (1–3). As demonstrated in a multicenter study encompassing 64 low- and middle-income countries, neonatal deaths contributed to 53.1% of the overall under-5 mortality (4). In 2023, 2.3 million children died within their first month of life worldwide, with an average of 6,300 deaths per day (5). In the same year, the global neonatal mortality rate was 17 per 1,000 live births (6), with Sub-Saharan Africa accountings for approximately 46%, of neonatal deaths (7).

A large proportion of these deaths are preventable. Evidence-based interventions have demonstrated potential in preventing up to 58% of deaths due to prematurity, 79% of intrapartum-related deaths, and 84% of infection-related deaths, all amongst the leading causes of neonatal mortality (8). Projections suggest that by 2025, such improvements may prevent approximately 71% of neonatal deaths, 33% of stillbirths, and 54% of maternal deaths annually (9). Additional research corroborates these findings. The World Health Organization (WHO) emphasizes that simple, cost-effective strategies such as skilled birth attendant, essential newborn care, infection prevention, and timely management of complications could drastically reduce neonatal mortality worldwide (10). Similarly, Lawn and al. (2014) highlight that nearly three-quarters of neonatal deaths could be prevented with universal access to quality care at birth and during the first week of life (11). More recent analyses reaffirm the critical role of integrated maternal and newborn health interventions for child survival. For instance, Hug and al. (2019) estimate that 2.4 million neonates died in 2019, mostly from preventable causes, and call for accelerated investment in proven interventions (12).

Mali is among the countries with the highest neonatal mortality in the world and still experiences significant challenges in improving child health outcomes. According to the Demographic Health Survey (DHS), Mali’s neonatal mortality rate (NMR) was 29 deaths per 1,000 live births in 2023-2024, contributing significantly to under-five mortality estimated at 87 deaths per 1,000 live births (3). Despite efforts to improve maternal and child health services, progress in reducing neonatal deaths has been slow, particularly in rural and underserved areas. The factors contributing to this neonatal mortality include inadequate healthcare access, poor maternal health, infectious diseases, and malnutrition among others (3).

However, improvements in service utilization and coverage do not necessarily translate into equivalent gains in neonatal survival, particularly when the quality, timeliness, and effectiveness of care remain inadequate.

The Sustainable Development Goals (SDGs) target of a reduction in neonatal mortality to 12 deaths per 1,000 live births by 2030 is unlikely to be achieved in Mali without major acceleration in the pace of mortality decline, which requires rapid scaling-up of effective and quality health interventions. To achieve this, identifying high-risk mortality areas and analyzing the spatial distribution of neonatal deaths are key steps towards a more effective allocation of health resources. Geographic disparities, healthcare access, and socio-economic factors strongly influence neonatal outcomes; analyzing the subnational spatiotemporal dynamics in neonatal mortality provides evidence for policy-making in addressing newborn health challenges.

In the context of maternal and child health, predictive modelling has proven valuable for predicting neonatal mortality, assessing the influence of socioeconomic and environmental factors, and identifying geographic clusters of vulnerability (13–16). These are planning and analytical tools that identify high-risk mortality areas, forecast future trends, and evaluate the potential impact of interventions (17,18). For example, spatial predictive models have been successfully used in sub-Saharan Africa to map out neonatal mortality hotspots and inform targeted public health responses(19–22).

Building on previous evidence of inequalities in neonatal mortality in Mali (23,24), we examined their spatiotemporal distribution and contextual determinants. Evidence remains limited on how these inequalities evolve, where high-mortality clusters persist, and how alternative intervention scale-up scenarios could alter subnational trends. We therefore analyzed neonatal mortality dynamics from 2012 to 2023, identified persistent high-mortality regions and associated factors, and projected potential reductions through 2035 under alternative scale-up scenarios for high-impact interventions.

## I. Methods

### 1. Study conceptual framework

We designed this study based on two conceptual and methodological frameworks which are the Mosley and Chen’s framework (1984) and the Lives Saved Tool (LiST) framework to analyze neonatal survival from a spatiotemporal perspective (25–27). The Mosley and Chen framework conceptualizes neonatal mortality as the outcome of dynamic interactions between socioeconomic, environmental, maternal, health service, and contextual determinants, as well as proximate biological and behavioral factors. Within this framework, environmental conditions, health service use, fertility-related factors, socioeconomic characteristics, and contextual influences are spatially distributed and temporally evolving, shaping differential access to and utilization of key maternal and neonatal health interventions. These interventions include preconception and family planning services, antenatal care, intrapartum and postpartum care, and neonatal preventive and curative care. They act on proximate biological and behavioral risk factors such as maternal anemia, preterm birth, small-for-gestational-age births, and neonatal infections.

The LiST framework complements the Mosley and Chen conceptual framework by linking changes in the coverage of evidence-based maternal and newborn health interventions to changes in mortality outcomes. Both frameworks together provide comprehensive insights for assessing the effects and mechanisms of spatiotemporal disparities and targeted interventions on neonatal mortality, accounting for regional heterogeneity and temporal trends in intervention coverage and risk factors. The frameworks also shaped the methodology and analytical approach of the study (Fig1).

**SGA** : Percent of children born in one of four categories: Preterm and small for gestational age (SGA), preterm and appropriate for gestational age (AGA), term and SGA, and term and AGA. SGA is defined as <10th percentile; preterm is defined as <37 weeks

### 2. Sources of data, study design, population

We used household survey data from the latest three Mali Demographic and Health Surveys (MDHS) conducted in 2012-2013, 2018, and 2023-2024. Gao, Kidal and Tombouctou regions were not surveyed during the 2012-2013 Demographic and Health Survey due to persistent conflicts and security issues resulting in missing data for these regions. The study population included all live births reported during the five years preceding each survey among women aged 15–49 years.

A two-stage cluster sampling design representative at national and sub-national (region) levels was used for the three surveys. In the first stage, enumeration areas (EAs) serving as primary sampling units were selected within urban and rural strata using probability proportional to size. In the second stage, households were systematically selected within each EA (7,15,28). The total number of enumeration areas (EAs) or clusters were 415, 345 and 407 in 2012, 2018 and 2023, respectively. Overall, 10,424 women were interviewed in 2012, 10,519 in 2018, and 17,231 in 202,3 for a total population of 38,174 women of reproductive age (15-49 years) across the three sources.

For the purpose of this study, the data were accessed on October 25^th^ 2025.

### 3. Variables

The unit of analysis of the study was a live birth occurring within the five years preceding the survey while the primary outcome was neonatal death (binary coded as 1 if the neonate died and 0 if neonate survived), defined as the death of a newborn within the first 28 days of life. The three datasets include a total number of 35722 live births and 2345 neonatal deaths. The study covariates comprised health service use determinants (numbers of ANC visits, tetanus vaccine at birth, skilled birth attendant, facility delivery, postnatal service), socio-economic and individual determinants (maternal age, parity, education, wealth quintile, occupation, marital status, child’s sex, birth interval, birth weight), contextual determinants (region, area of residence) and environmental and WASH variables (unimproved water source, unimproved toilet, hand washing).

### 4. Interventions

Table 1 summarizes the interventions for neonatal mortality reduction included in the Lives Saved Tool (LiST) framework, grouped under three domains: prevention, immediate newborn and postnatal care and maternal interventions during pregnancy and delivery.

**Table 1:**
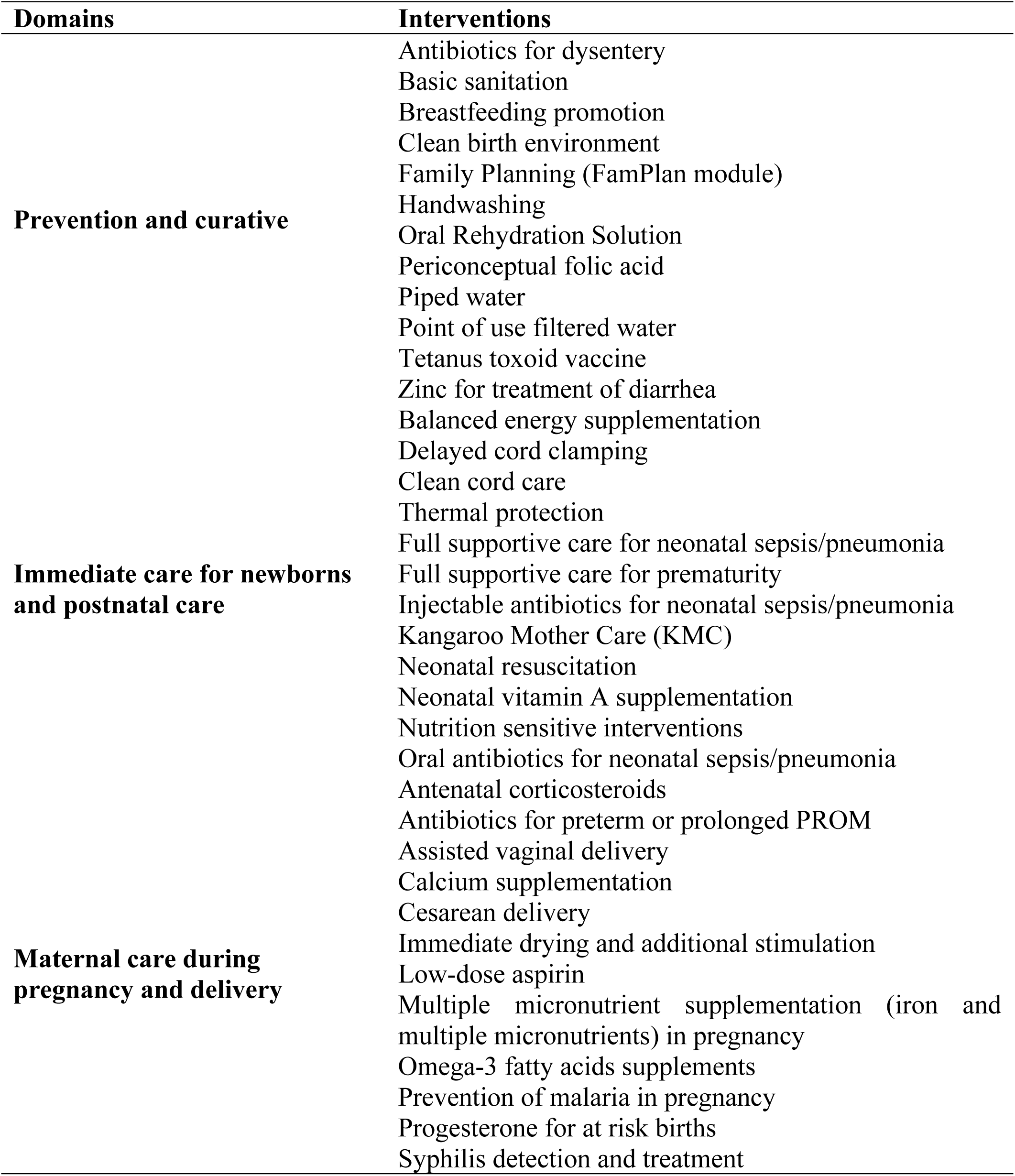
Interventions for neonatal mortality reduction included in the Lives Saved Tool (LiST) framework.

### 5. Data analysis Statistical analysis

We merged the three datasets and used Stata version 19.5 for analysis. All analyses accounted for the complex survey design, including stratification, clustering, and sampling weights. We first conducted descriptive and exploratory analyses to characterize trends in neonatal mortality across the three survey rounds.

We mapped weighted neonatal mortality rates using Magrit to visualize spatial and temporal patterns across survey years (29). To facilitate comparability between surveys, we classified mortality rates using the equal-interval discretization method (30).

We fitted multilevel mixed-effects logistic regression models to identify individual, household and community level factors associated with neonatal mortality by using “melogit” code of STATA (22,31). The intraclass correlation coefficient (ICC) was estimated to quantify the proportion of variance attributable to the community level (9). We assessed the Global and local spatial autocorrelation using Moran’s I statistic computed from Geoda (2,21,32,33). All spatial analyses were conducted using QGIS version 3.40.15.

Multicollinearity was assessed using Variance Inflation Factors (VIF). All VIF values were below 3 (mean VIF = 2.47), indicating no evidence of problematic multicollinearity among explanatory variables. Because substantial missing data reduced the analytical sample, a parsimonious multivariable model was retained to maximize statistical power and minimize selection bias.

#### Geospatial modelling

The spatial analysis aimed to describe cluster and regional variation in neonatal mortality and was conducted using QGIS version 3.40.15. We carried out the analysis using administrative regions as analytical cluster aggregating neonatal mortality data. We constructed a spatial contiguity matrix based on a neighborhood criterion (queen contiguity), whereby regions sharing a common boundary were defined as neighbors.

To examine the spatial distribution of neonatal mortality across regions, we performed a global spatial autocorrelation analysis using Moran’s I statistic in GeoDa (31). This method assesses whether the observed distribution exhibits a statistically significant spatial structure at the regional level. A positive and statistically significant Moran’s I value indicates that regions with similar levels of neonatal mortality tend to cluster geographically. Local Indicators of Spatial Association (LISA) were subsequently used to identify the location and type of local spatial clusters, including high-high (hotspot) and low-low (coldspot) clusters.

#### LiST modelling

We used the Lives Saved Tool (LiST) to estimate the potential impact of scaling up maternal and newborn health interventions on neonatal mortality at the national and subnational levels (34). LiST is a mathematical modelling tool that links changes in intervention coverage to changes in mortality outcomes and enables the assessment of alternative programmatic scenarios (27,35–38). Consistent with its application in maternal and child health program evaluation, the LiST tool helped to estimate the number of neonatal deaths that could be averted under different coverage scale-up scenarios and to identify interventions with the greatest potential impact on neonatal survival.

We developed simulation scenarios to project the number of neonatal lives that could be saved by increasing the coverage of selected interventions by 2035. We selected interventions based on their relevance for preventing neonatal deaths in Mali and their availability within the LiST framework. The interventions included skilled birth attendant (SBA), facility delivery, caesarean section (CS), intermittent preventive treatment of malaria during pregnancy (IPTp), kangaroo mother care (KMC), neonatal resuscitation, antibiotics for neonatal sepsis, screening and treatment of maternal infections, multiple micronutrient supplementation, and early and exclusive breastfeeding.

Baseline intervention coverage estimates for 2026 were derived primarily from the 2023 MDHS; for interventions not measured in the MDHS, default Mali-specific LiST values were retained (**S1Table**). Because Mali had not adopted a new national health strategic plan following the expiration of the Social and Healthy Development decadal Plan (PDDSS) in 2023, no official coverage targets were available for projections to 2035. Coverage trajectories were therefore projected using linear extrapolation of trends observed across the 2012–2023 MDHS rounds, and two additional accelerated scale-up scenarios were modeled by increasing the projected slope by 20% and 40%, respectively, with coverage capped at 100%. Selected interventions included skilled birth attendant, facility delivery, cesarean section, IPTp, Kangaroo Mother Care, neonatal resuscitation, antibiotics for neonatal sepsis, screening and treatment of maternal infections, multiple micronutrient supplementation, and early and exclusive breastfeeding. Several analyses using LiST, including those conducted in Mali under the National Evaluation Platform (NEP) or in other low- and middle-income countries, have applied a similar structure to assess the achievability of national targets and estimate the impact of interventions actually implemented (39–42).

Three projection scenarios were modelled and validated through a national expert consultation involving representatives from INSP, the Planning and Statistics Unit, the Directorate-General for Health and Public Hygiene, and the the Gobal Financing Facility (GFF) country coordinator. Scenario assumptions were assessed based on historical intervention coverage trends (2012–2023 MDHS), previous national health priorities and targets, experts’ program implementation experience, and the feasibility of achieving projected coverage levels by 2035.

#### Projection 1: Linear Trend

Projection 1 extrapolated the annual absolute changes in intervention coverage observed between the 2012 and 2023 DHS surveys through 2035. In the absence of updated national targets, this conservative scenario serves as the reference trajectory for assessing the incremental impact of accelerated scale-up strategies. The linear projections followed standard LiST modelling approaches (27,43).

#### Projection 2: Moderate Scale-Up (+20% of slope)

Projection 2 increased coverage levels from slope by 20 percent by 2035. This scenario reflects achievable improvements through strengthened implementation, better service delivery, and moderate increases in resource allocation. It assumes programmatic acceleration without major systemic reform. Projection 2 estimates the potential reduction in neonatal mortality under realistic expansion conditions and represents a feasible pathway toward improved maternal and newborn health outcomes. Moderate and accelerated scale-up scenarios were modeled in line with previous LiST-based analyses assessing potential lives saved under expanded coverage (35,44,45).

#### Projection 3: Accelerated Scale-Up (+40% of slope)

Projection 3 was designed to estimate the potential reduction in neonatal mortality under an ambitious acceleration. Coverage levels increased by 40 percent, capped at 100%, if needed by 2035. This increase was selected to represent an ambitious but plausible acceleration scenario rather than a predefined requirement for achieving the SDGs. This threshold is consistent with acceleration scenarios commonly used in LiST-based modeling studies (46,47)

This scenario assumes strong political commitment, sustained investment, and comprehensive health system strengthening. It represents an ambitious scale-up pathway and provides an upper-bound estimate of the potential reduction in neonatal mortality that could be achieved under accelerated intervention coverage(48,49).

## Ethical approval

Ethical approval was not required for this study based on publicly available, de-identified secondary data from the Demographic and Health Surveys Program.

## Results

### Temporal and regional variation in neonatal mortality

The national neonatal mortality rates decreased from 34 deaths per 1,000 live births (95% CI: 31–38) in 2012 to 32 per 1,000 live births (95% CI: 29–36) in 2018, and to 29 per 1,000 live births (95% CI: 26–32) in 2023. Temporal and regional variation in neonatal mortality across the three survey rounds is shown Figure 2.

**Figure 1:**
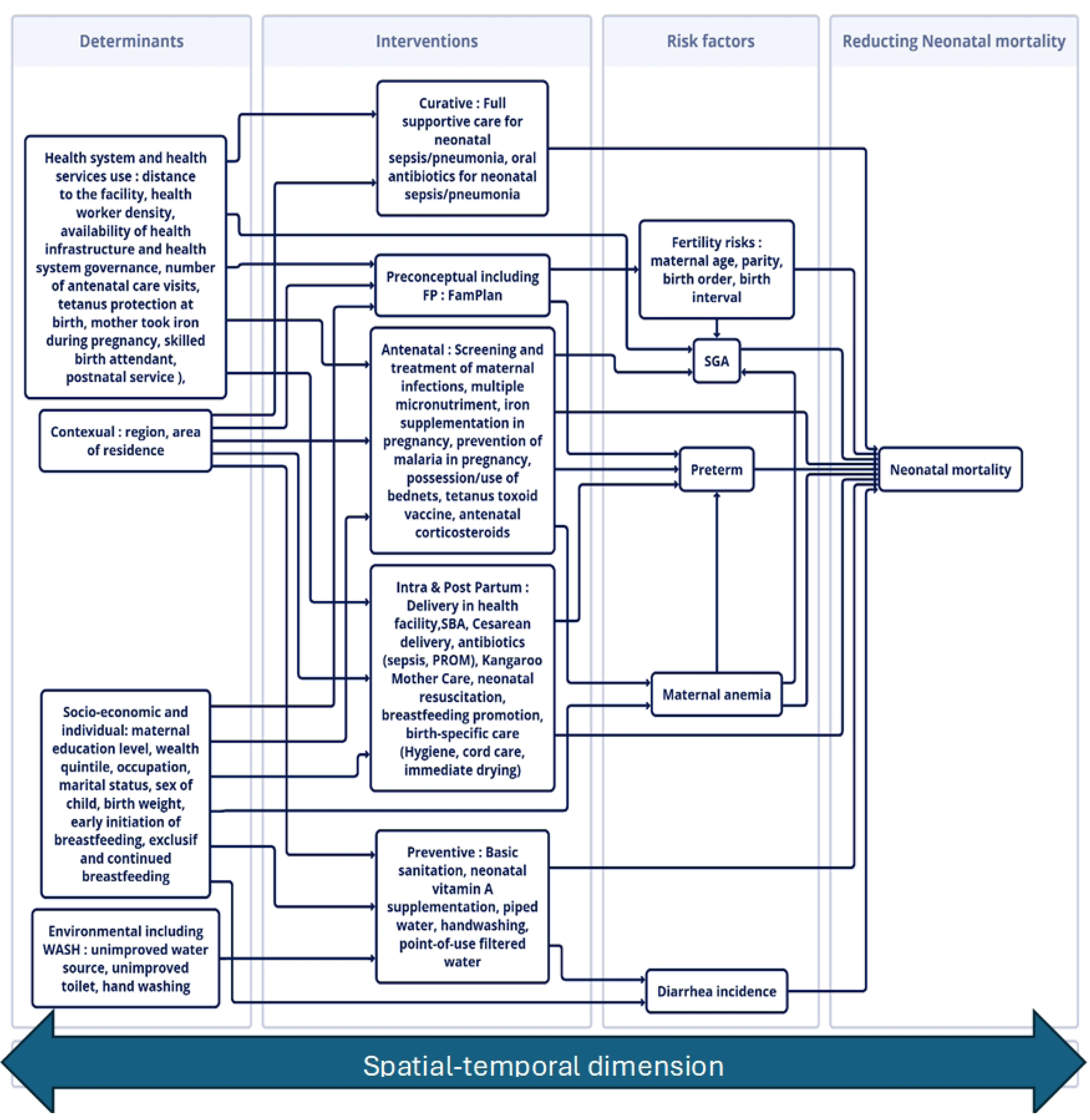
Conceptual framework for reducing neonatal mortality in Mali, adapted from Mosley and Chen (1984) and the Lives Saved Tool (LiST) frameworks.

**Figure 2:**
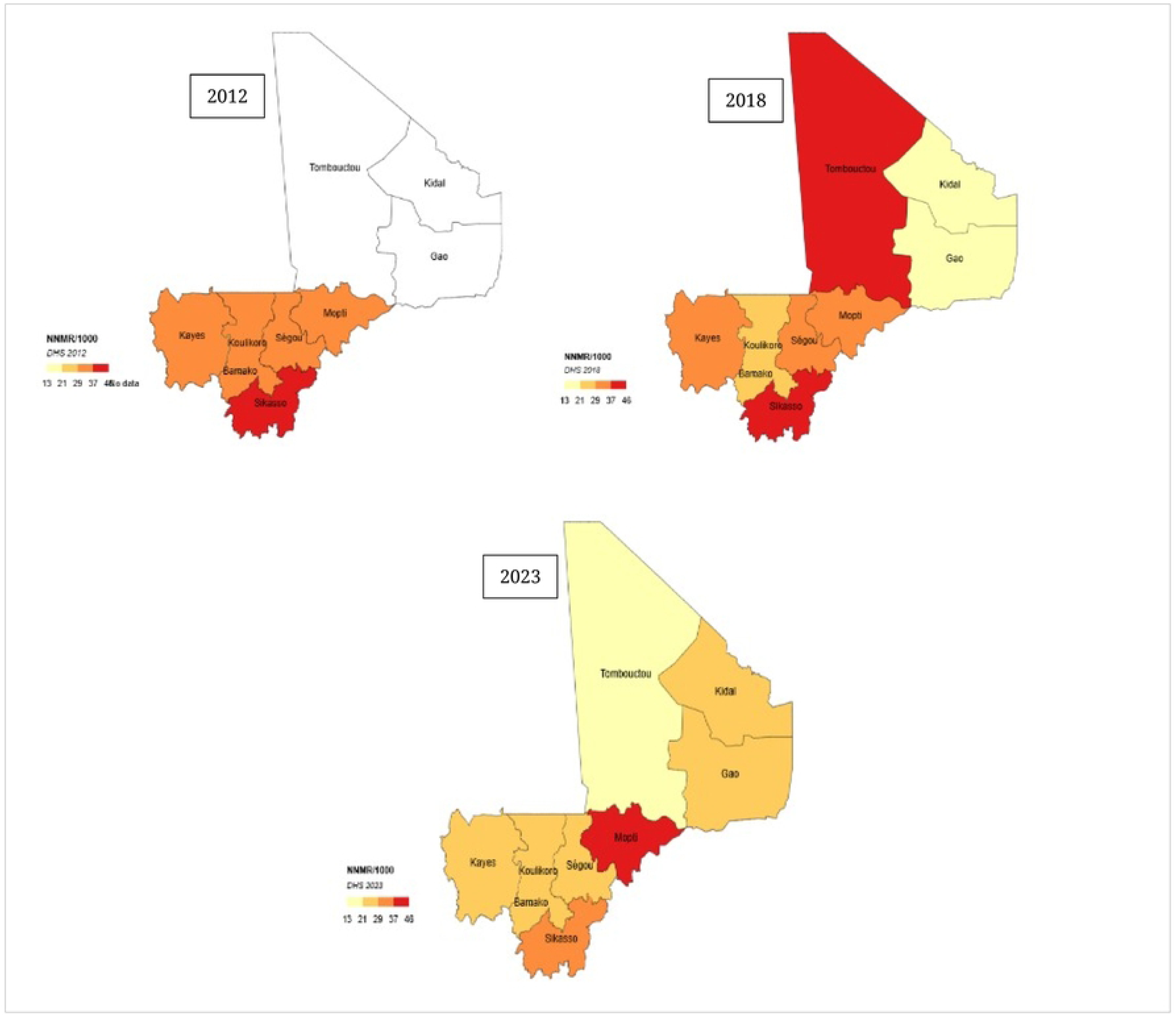
Regional trends of empirical neonatal mortality rates from 2012 to 2023

In 2012, Sikasso had the highest neonatal mortality rate (44.2 per 1,000 live births), followed by Kayes (35.2), whereas Bamako had the lowest rate (24.4). In 2018, the highest neonatal mortality rate was in Tombouctou (45.5 per 1,000 live births) and remained high in Sikasso (41.8 per 1,000 live births), Kayes (36.1 per 1,000 live births), Ségou (34.6 per 1,000 live births), and Mopti (34.2-per 1,000 live births). In 2023, Mopti became the region with the highest neonatal mortality rate (37.8 per 1,000 live births), followed by Sikasso (32.1 per 1,000 live births). Tombouctou showed the largest decline, from 45.5 in 2018 to 18.7 per 1,000 live births in 2023. Although neonatal mortality declined substantially in Sikasso (44.2 to 32.1 per 1,000 live births) and Ségou (34.6 to 28.6 per 1,000 live births between 2018 and 2023), these regions, together with Mopti, consistently remained among those with the highest neonatal mortality rates during the study period.

Maps display regional neonatal mortality rates estimated from DHS 2012, DHS 2018, and DHS 2023.

### Neonatal mortality rates by sociodemographic and reproductive characteristics

The supplementary *S2* Table presents neonatal mortality according to child, maternal, household, and regional characteristics across the three DHS surveys. Overall, neonatal mortality declined from 34 to 29 deaths per 1,000 live births between 2012 and 2023. Mortality remained consistently higher among male neonates, first births, births spaced by less than two years, children of mothers younger than 20 years, and rural populations. Regional differences were observed across survey rounds. In 2018, neonatal mortality was significantly higher in Ségou than in Bamako (OR 2.19, 95% CI: 1.15–4.20), while in 2023–2024, significantly higher odds of neonatal mortality were observed in Sikasso (OR 2.06, 95% CI: 1.53–2.78), Ségou (OR 2.13, 95% CI: 1.25–3.63), and Mopti (OR 2.86, 95% CI: 1.44–5.67), compared with Bamako.

### Global spatial autocorrelation of neonatal mortality

Global Moran’s I statistics showed positive and statistically significant spatial autocorrelation across all survey years (p < 0.001), indicating the presence of spatial clustering in neonatal mortality (Table 2). Moran’s, I increased from 0.144 in 2012 to 0.243 in 2018 and was 0.172 in 2023, indicating positive spatial dependence in the geographic distribution of neonatal mortality throughout the study period.

**Table 2:** Testing for global spatial auto correlation using Moran’s I statistics.

| Survey year | Moran's I | P-value |
| --- | --- | --- |
| 2012 | 0.144 | <0.001 |
| 2018 | 0.243 | <0.001 |
| 2023 | 0.172 | <0.001 |

*Note: Positive Moran's I values indicate positive spatial autocorrelation, P-values <0.05 indicate statistically significant spatial autocorrelation.*

### Multilevel analysis of neonatal mortality

The null multilevel model yielded an Intraclass correlation (ICC) of 10.3% (95% CI: 6.1%– 16.8%), indicating that approximately one-tenth of the variation in neonatal mortality was attributable to differences between communities.

### Spatial autocorrelation and clustering of neonatal mortality

Figure 3 below presents the Local Indicators of Spatial Association (LISA) cluster maps of neonatal mortality across the three survey rounds. Five spatial patterns were identified: 1) areas with no statistically significant spatial autocorrelation; 2) high-high clusters (hot spots), where high values were surrounded by high neighboring clusters with similarly high mortality; 3) low-low clusters (cold spots), where low values were surrounded by neighboring low-mortality clusters; 4) low-high spatial outliers; and 5) high-low spatial outliers.

**Figure 3:**
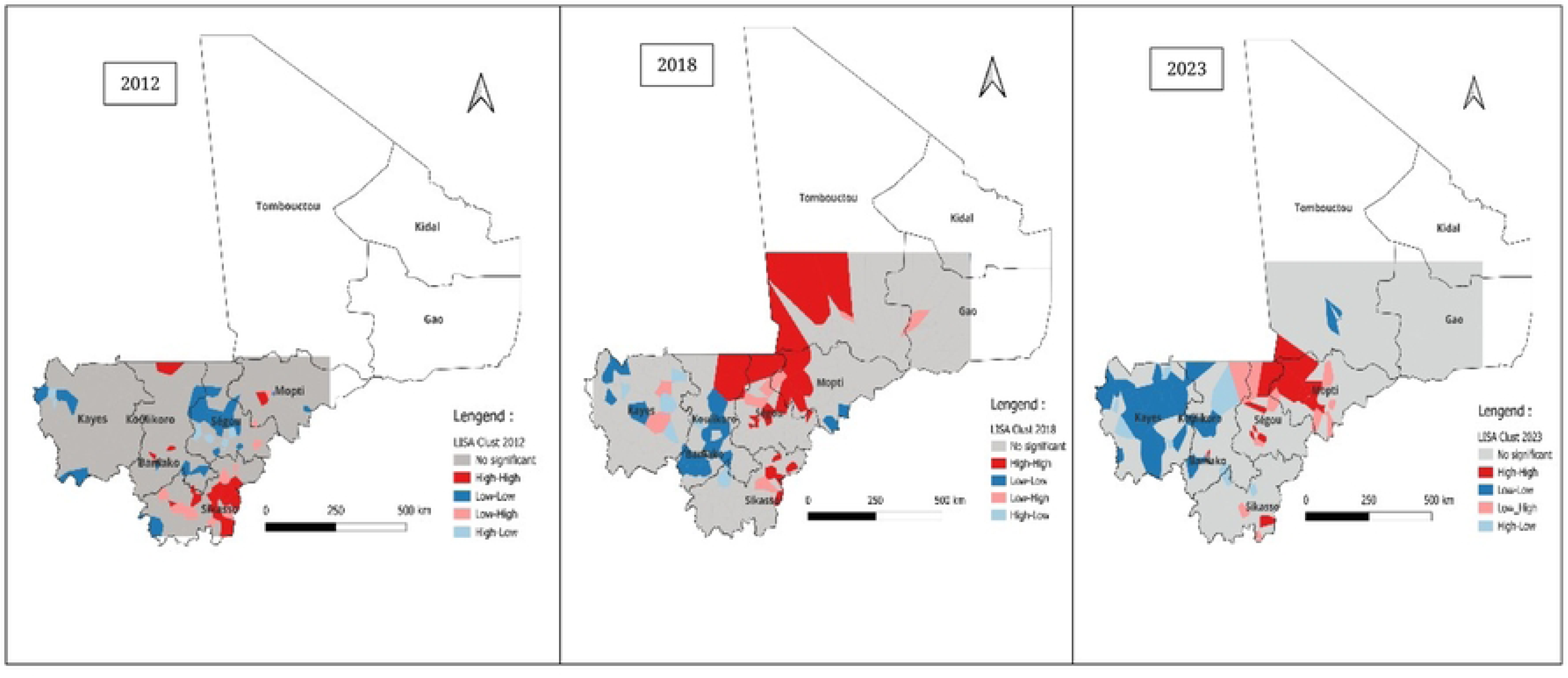
Geographical variation of neonatal mortality rate of Mali, from MDHS V, VI, VII

In 2012 significant high–high clusters (hotspots) were concentrated in Sikasso and Mopti. Low– low clusters (cold spots) appear mainly in the southwestern regions. A few low–high and high– low spatial outliers were observed in central areas. As noted previously, no data were available for the northern regions (Tombouctou, Gao and Kidal), which were not covered by the 2012 survey.

In 2018, high–high clusters were observed in Sikasso, Mopti and Ségou, with an additional high–high cluster in Tombouctou. Low–low clusters remained concentrated in the western regions, particularly Kayes and Koulikoro. A limited number of high–low and low–high spatial outliers were observed in central and southern Mali.

In 2023, high–high clusters remained concentrated in Mopti, Sikasso and parts of Ségou, although their spatial extent was reduced compared with 2018. Low–low clusters persisted in the western regions, especially in Kayes and Koulikoro. Only a limited number of spatial outliers were identified in central and southern Mali.

High-high clusters indicate hotspot regions, while low-low clusters indicate cold spots.

### Factors associated with neonatal mortality

The supplementary S3 Table presents the results of the multilevel logistic regression analyses for the three DHS surveys. The magnitude and statistical significance of the associations varied across survey rounds. Male sex and a short birth interval were consistently associated with higher odds of neonatal mortality across the three survey rounds, although the magnitude of the associations varied over time. Postnatal care utilization was associated with lower odds of neonatal mortality in 2012 and 2018, but this association was no longer statistically significant in 2023. Similarly, improved sanitation was associated with lower odds of neonatal mortality in 2012 and 2018 but not in 2023.

In 2023, regional disparities became more pronounced, with significantly higher odds of neonatal mortality in Sikasso, Ségou and Mopti compared with Bamako. Household wealth was not associated with neonatal mortality in earlier surveys but became significant in 2023, with higher odds among children from richer households.

### Projected impact of scaling up maternal and newborn health interventions on neonatal mortality

Figure 4 presents the projected trends in neonatal mortality rates in Mali from 2026 to 2035 under three different LiST projection scenarios. All scenarios showed a progressive decline in neonatal mortality over the period.

**Figure 4:**
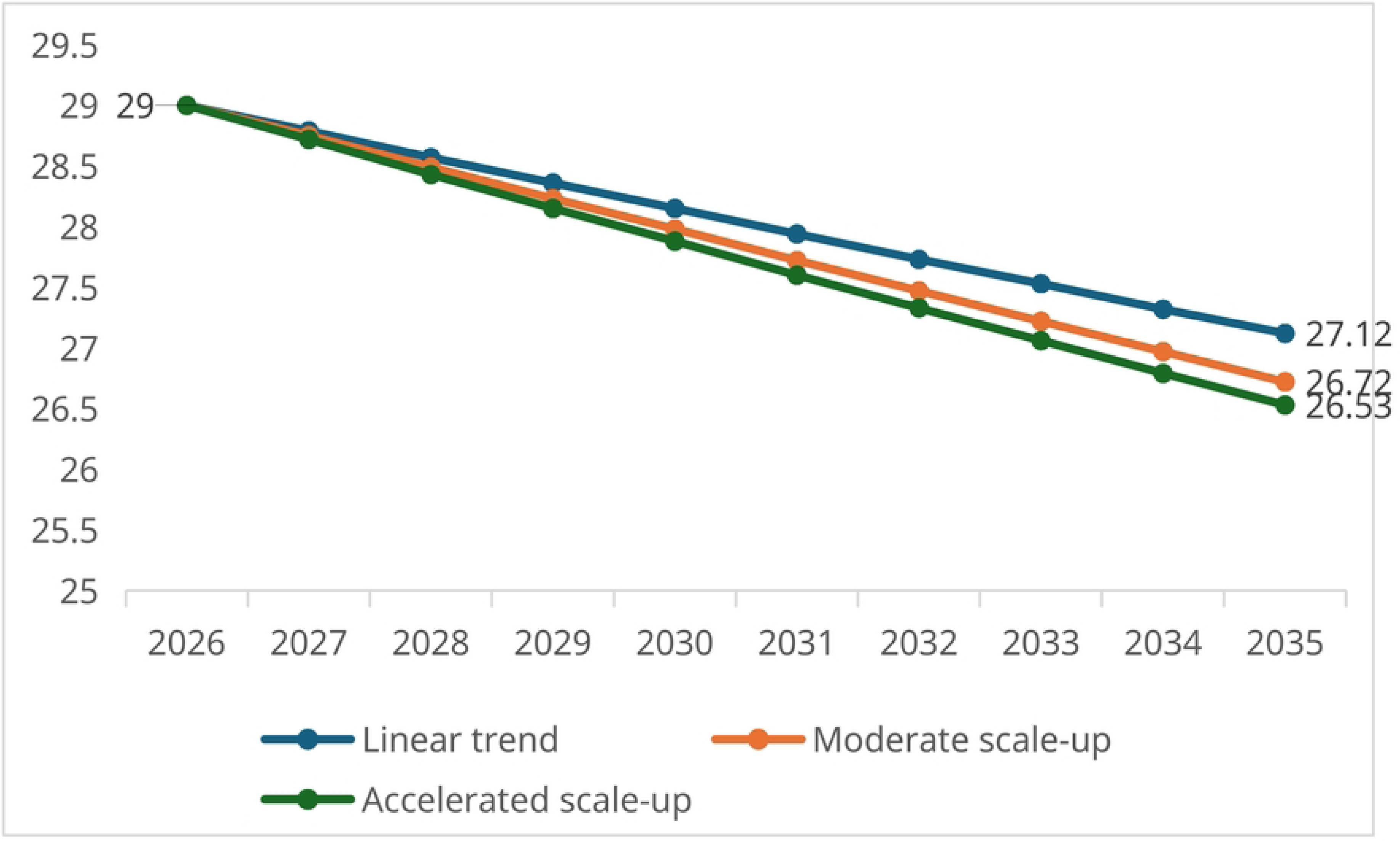
Projected neonatal mortality rate from 2026 to 2035 in Mali. based on three different LiST projections

Projection 1: Linear trend. Projection 2: Moderate scale-up. Projection 3: Accelerated scale up.

Under Projection 1 (linear trend projection), the neonatal mortality rate was projected to decline from 29 deaths per 1,000 live births in 2026 to 27.12 deaths per 1,000 live births in 2035, corresponding to an estimated 10,218 neonatal lives saved during the period.

Projection 2 showed a greater reduction, with neonatal mortality decreasing from 29 to 26.72 deaths per 1,000 live births between 2026 and 2035, representing approximately 12,333 neonatal lives saved. Similarly, projection 3 showed the largest reduction in neonatal mortality, decreasing from 29 to 26.53 deaths per 1,000 live births by 2035, with an estimated 13,463 neonatal lives saved (Table 3).

**Table 3:**
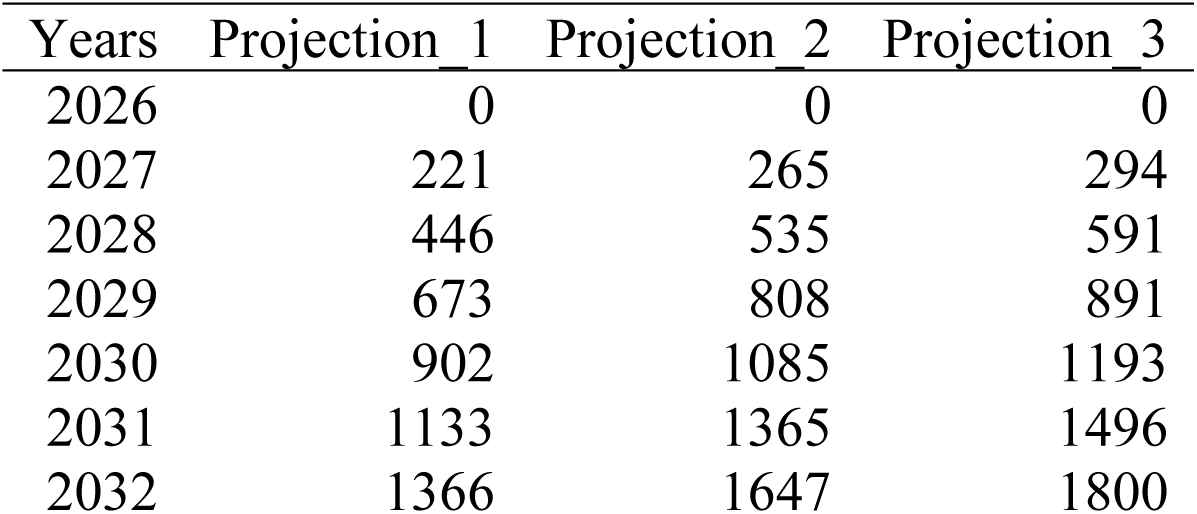
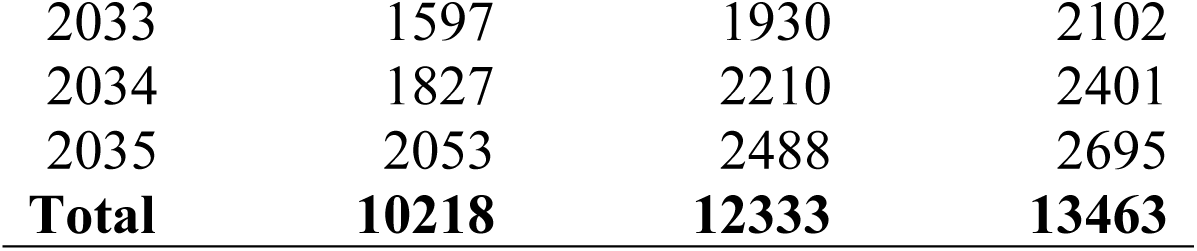
Number of lives saved from 2026 to 2035 in Mali based on three different LiST projections.

The figure 5 below shows the distribution of neonatal lives saved by intervention under each of the three scenarios. Of the various interventions modelled, six are responsible for the largest number of lives saved (Fig 5). The top three interventions in projections 2 and 3 (thermal protection, neonatal resuscitation, clean cord care) together were responsible for 53.29% and 53.57 % of neonatal lives saved, respectively, in each projection.

**Figure 5:**
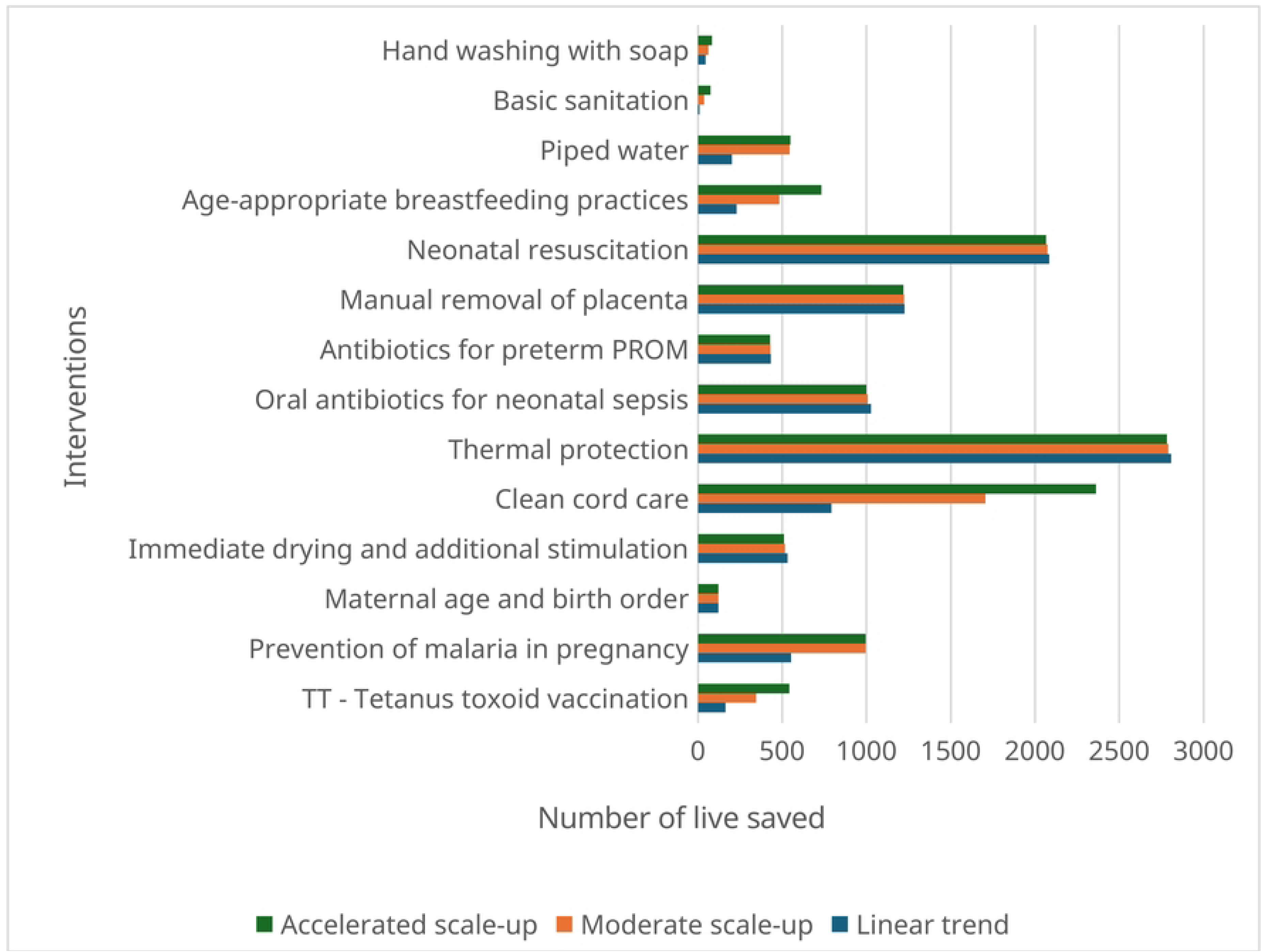
Projected neonatal of lives saved from 2026 to 2035 in Mali by intervention, based on three different LiST projection

## Discussion

This study aimed to identify geospatial clustering of the risk of neonatal deaths at the regional and MDHS cluster level to identify driving interventions and high-risk regions. We used the Lives Saved Tool (LiST), a modelling tool bringing together evidence-based health interventions, interventions from country health policy monitoring and evaluation frameworks, and national priority programmatic interventions.

Our findings revealed substantial and persistent geographic inequalities in neonatal mortality, and varying effects of maternal, neonatal, and healthcare related factors over time. At the national level, neonatal mortality declined only modestly over the study period, while important regional disparities persisted and some determinants changed in magnitude and statistical significance across survey rounds. In addition, the LiST projections suggested that the current pace of progress may be insufficient for Mali to achieve expected neonatal mortality reduction targets without accelerated improvements in maternal and newborn health interventions. To our knowledge, this is one of the first studies in Mali to combine spatial analysis, multilevel modelling, and LiST projections to simultaneously identify current geographical inequalities, determinants of neonatal mortality, and the potential impact of scaling up evidence-based interventions through 2035. This integrated approach provides evidence that can support geographically targeted and evidence-informed health planning.

The global Moran’s I statistics confirmed that neonatal mortality was not randomly distributed, indicating significant spatial autocorrelation between neighboring clusters. The LISA analysis further identified distinct high–high and low-low clusters, showing important spatial heterogeneity in neonatal mortality patterns over time. Persistent high–high clusters were mainly observed in the regions in the central part of the country. These findings are consistent with studies from several sub-Saharan African countries reporting marked geographical clustering and spatial inequalities in neonatal mortality. In Ethiopia, significant spatial autocorrelation and high-mortality clusters have been documented across successive DHS rounds, while a recent study from Nigeria similarly identified geographically clustered patterns of neonatal mortality using Global Moran’s I and LISA analyses (19,20).

These findings suggest the presence of geographically concentrated risk factors affecting neonatal survival. In Mali, these geographical inequalities may be further exacerbated by insecurity, which has disrupted health-service delivery, damaged or constrained health infrastructure, and limited access to essential maternal and newborn health services, particularly in the northern and central regions (50–52). Geographic inequalities in neonatal survival have also been reported in fragile settings in Somalia (53). However, insecurity alone is unlikely to explain the observed spatial pattern. Geographic disadvantage may also reflect long travel distances to health facilities, weak referral and transport systems, shortages or uneven distribution of skilled health personnel, limited availability of functional newborn care facilities and essential commodities, poverty, seasonal barriers to access, and population displacement. The national strategies should prioritize geographically targeted investments in conflict-affected regions, while also strengthening referral networks, transport systems, human-resource availability, facility readiness, and continuity of care across the continuum from birth to the early postnatal period.

The multilevel regression analysis identified several factors significantly associated with neonatal mortality, although the strength and significance of these associations changed over time. In 2012, male newborns had significantly higher odds of neonatal mortality compared with female newborns. The higher mortality observed among male newborns is consistent with previous studies (54,55). Longer birth intervals, maternal age ≥20 years, tetanus vaccination during pregnancy, postnatal care utilization, and improved sanitation were associated with lower odds of neonatal mortality. The association between longer birth intervals and lower neonatal mortality is consistent with previous evidence from sub-Saharan Africa and other low- and middle-income countries, where short birth intervals have been associated with increased risks of neonatal and perinatal mortality (56–58).

In 2018, birth intervals of three years, postnatal care utilization, and improved sanitation were associated with lower odds of neonatal mortality. However, several maternal and healthcare related factors that were significant in 2012, including maternal age and tetanus vaccination, were no longer statistically significant. This attenuation may partly reflect improvements in maternal and child health service coverage and utilization between 2012 and 2018, particularly facility based delivery, postnatal care, and antenatal care, despite persistent regional and socioeconomic inequalities (52,59,60).

By 2023, regional remained important determinants of neonatal mortality, with mortality significantly higher in Sikasso, Ségou, and Mopti compared with Bamako. The analysis also showed an unexpected pattern in the association between household wealth and neonatal mortality. However, this finding should be interpreted with caution because the relatively small number of observations, combined with the inclusion of multiple covariates in the multivariable model, resulted in limited statistical power to estimate this association precisely. Several inequalities observed in earlier surveys, including those related to birth interval, maternal age, tetanus vaccination, postnatal care, and sanitation, became less pronounced over time, although the observed changes should be interpreted as changes in estimated associations rather than definitive evidence that inequalities in effective care had disappeared.

In contrast, the association between neonatal sex and mortality persisted across surveys, suggesting that this difference may not be readily influenced by overall improvements in maternal and newborn healthcare coverage and may instead reflect underlying biological or clinical vulnerabilities that require further investigation. Regional disparities also persisted, highlighting the continued influence of geographical inequalities on neonatal survival.

Despite the achievements in maternal and child health service coverage over the last decade in Mali, neonatal mortality has declined relatively slowly compared to those of the under-five (40). This persistent gap between improvements in service coverage and neonatal mortality outcomes highlights the challenges of translating increased coverage into effective reductions in neonatal deaths. The failure to achieve the neonatal mortality target established in the 2014– 2023 National Health Development Plan (PDDSS) further illustrates these implementation challenges. Although national initiatives such as results-based financing (RBF), the creation of national office of reproductive health, and community health programs may contribute to improving maternal and newborn outcomes, our findings suggest that stronger, geographically targeted, and quality focused strategies will be required to substantially accelerate neonatal mortality reduction over the coming decade.

The LiST projection analysis showed that neonatal mortality in Mali is expected to decline gradually between 2026 and 2035 under all projection scenarios. However, the projected reductions remain insufficient to achieve substantial acceleration in neonatal survival if intervention coverage continues to improve at the current pace. Under the linear trend scenario, neonatal mortality would decline only modestly from 29 to 27.12 deaths per 1,000 live births by 2035, whereas more ambitious scenarios assuming greater improvements in maternal, newborn, child health and nutrition (MNCH&N) intervention coverage produced larger reductions in neonatal mortality and higher numbers of neonatal lives saved.

Our findings are consistent with previous national reports showing that neonatal mortality has declined more slowly than under-five mortality in Mali despite improvements in maternal and child health service coverage (40). Although important progress has been achieved in maternal and child health service coverage over the last decade, neonatal mortality has declined more slowly. This persistent gap between improvements in service coverage and neonatal mortality outcomes highlights the challenges of translating increased coverage into effective reductions in neonatal deaths. The inability of Mali to achieve the neonatal mortality target established in the 2014–2023 National Health Development Plan (PDDSS) further illustrates these implementation challenges. Although national initiatives such as results-based financing (RBF), creation of national office of reproductive health, and community health programs may contribute to improving maternal and newborn outcomes, our findings suggest that stronger, geographically targeted, and quality focused strategies will be required to substantially accelerate neonatal mortality reduction over the coming decade.

Similar LiST analyses from Tanzania and Ethiopia have shown the potential of scaling up high-impact maternal and newborn interventions to accelerate mortality. In Tanzania, LiST modelling of small and sick newborn care highlighted the substantial potential gains from expanding coverage of key neonatal interventions, while analyses from Ethiopia similarly demonstrated the mortality reductions achievable through increased intervention coverage (61,62). Our projections extend this evidence to Mali, showing that accelerating coverage expansion could substantially reduce neonatal mortality, but that coverage gains alone may be insufficient to overcome persistent quality of care and geographical disparities.

With regard to interventions, in all scenarios, a relatively small number of interventions were sufficient to save the majority of neonatal lives. Thermal care, neonatal resuscitation, and clean cord care emerged as the most influential interventions, together contributing 53.29% and 53.57% of all neonatal lives saved under Projections 2 and 3, respectively. These findings are consistent with previous evidence showing thermal care and neonatal resuscitation directly address some of these leading causes of death, while clean cord care helps prevent neonatal infections, particularly sepsis, which remains a major challenge in sub-Saharan Africa (63,64). The large contribution of these interventions highlights the importance of improving care during childbirth and the first days of life, when most neonatal deaths occur.

The concentration of projected lives saved among a limited number of high-impact interventions, suggest that Mali could maximize the impact of limited health resources by prioritizing high-impact newborn interventions within the national SRMNIA-Nut strategy, particularly in underserved and conflict-affected regions. Expanding training on neonatal resuscitation, ensuring the availability of thermal care materials and practice, and strengthening clean cord care practice should therefore constitute key programmatic priorities.

Beyond identifying determinants of neonatal mortality, this study provides actionable evidence for health planning in Mali. The findings support a combination of nationwide improvements in service coverage and geographically targeted interventions in high-burden and conflict-affected areas. Prioritizing high-impact interventions, including neonatal resuscitation, thermal protection, and clean cord care, while strengthening the quality of facility- and community-based newborn care, could accelerate progress towards the SDG neonatal mortality target.

Our analysis has some limitations. First, in the absence of an updated national strategic plan following the end of the most recent PDDSS in 2023, no officially endorsed coverage targets were available for projection to 2035, Consequently, coverage trends were extrapolated linearly, which may not fully reflect future programmatic changes or policy shifts. Second, Gao, Kidal and Tombouctou regions were not surveyed during the 2012 Demographic and Health Survey due to persistent conflicts and security issues resulting in missing data for these regions. Third, not all LiST intervention indicators were available in the DHS datasets. For interventions with unavailable indicators, default Mali-specific LiST values were used, which may have introduced some uncertainty into the projections. Despite these limitations, this study has several important strengths. It combines nationally representative DHS data collected over more than a decade with complementary analytical approaches including spatial statistics, multilevel modelling, and LiST projections. This triangulation allowed us to identify not only the geographical distribution and determinants of neonatal mortality but also to estimate the potential public health impact of scaling up evidence-based interventions. The integration of these approaches provides comprehensive evidence for priority setting and resource allocation in maternal and newborn health programs.

## Conclusion

This study demonstrates that neonatal mortality in Mali has declined only modestly while substantial spatial inequalities persist and the determinants of mortality have evolved over time. The most ambitious LiST scenario suggests that accelerated intervention scale-up could reduce mortality, but the projected 2035 level of approximately 26.5 deaths per 1,000 live births remains far above the SDG target of 12. These findings indicate that expanding intervention coverage alone will not be sufficient. Mali will need to combine targeted scale-up of high-impact interventions particularly thermal care, neonatal resuscitation, and clean cord care with improvements in effective coverage, quality of care, facility readiness, provider competency, referral systems, essential commodities, and continuity of early postnatal care. Priority should be given to regions and districts with persistent high mortality and constrained access to effective care, while strengthening district-level monitoring of both coverage and quality.

## Supporting information

S1 Table: Baseline and endline coverage levels for each of the three projections at national level (DOCX).

S2 Table: Neonatal l mortality rates by sociodemographic and reproductive characteristics, Mali DHS 2012–2023 (per 1,000 live births) (DOCX).

S3 Table: Multilevel mixed-effects logistic regression of factors associated with neonatal mortality in Mali, DHS 2012, 2018 and 2023 (DOCX).

S4 Table: Description of the sample and response rates in 2012, 2018 and 2023 MDHS (DOCX).

S1 Fig: List of interventions implemented in LiST. with risk factors and causes of death to reduce neonatal mortality ((TIF)).

## Acknowledgments

The authors acknowledge the Countdown to 2030 initiative for providing the fellowship opportunity that supported this work. We also acknowledge the African Population and Health Research Center (APHRC) for technical support and guidance throughout the study. We thank the National Public Health Institute (INSP), Mali, for its institutional support.

## Author Contributions

FBT wrote the first draft with substantial inputs from AM, CSS, YK, FS and MT contributed to study conceptualization. IT, MB, HD contributed to the methodology and formal analysis. FD and AM provided supervision. All authors contributed to manuscript review and editing and approved the final version.

## Data Availability

The datasets analyzed in this study were obtained from the Demographic and Health Surveys (DHS) Program. The Mali DHS datasets are available upon registration and application through the DHS Program website (https://dhsprogram.com/), subject to the DHS Program’s terms of data access and use.

## Competing Interests

The authors declare that they have no competing interests

## Funding

This study was conducted as part of the Countdown to 2030 Fellowship Program funded by the Bill and Melinda Gates Foundation through a grant to APHRC (Grant number INV-003416). The conclusions and opinions expressed in this work are those of the authors alone and shall not be attributed to the Foundation.

## Notes

### Competing Interest Statement

The authors have declared no competing interest.

### Clinical Trial

N/A

